# The Locus Coeruleus-Basal Forebrain tract in Lewy body disease: autonomic dysfunction and cognition

**DOI:** 10.64898/2026.08.03.26359591

**Authors:** Abhimanyu Mahajan, Kevin R Duque, Dong-Woo Ryu, Alberto J. Espay, Brady Williamson

## Abstract

**Background:** Cardiovascular autonomic dysfunction and cognition associate with each other throughout the disease course in Lewy body disease, and this relationship is poorly understood.

**Methods:** We performed a cross-sectional imaging analysis to investigate the locus coeruleus – basal forebrain tract. Deterministic tractography based on quantitative anisotropy was performed. Using Bayesian semiparametric proportional-odds modeling, we evaluated the association between neurogenic orthostatic hypotension and tract characteristics with adjustment for age at imaging, sex, and education.

**Results:** In the demographic-adjusted model, neurogenic orthostatic hypotension was associated with lower bilateral fourth-quarter volume (OR = 0.19; Pr[OR < 1] = 0.987), left fourth-quarter volume (OR = 0.13; Pr[OR < 1] = 0.997), left total surface area (OR = 0.19; Pr[OR < 1] = 0.990), and left total volume (OR = 0.22; Pr[OR < 1] = 0.982). These associations were directionally consistent and of similar magnitude after additional adjustment for cognitive severity, high blood pressure, disease duration, and combined high blood pressure and disease duration. Participants with impaired cognition showed lower bilateral fourth-quartervolume (OR = 0.33; Pr[OR < 1] = 0.964)

**Conclusion:** Neurogenic orthostatic hypotension is associated with lower locus coeruleus-basal forebrain tract volume. The inverse correlation between volume and cognitive impairment suggest a mechanistic relationship.

## Introduction

Cardiovascular autonomic dysfunction (CVAD), manifesting as orthostatic hypotension (OH) is documented in approximately 50% of patients with Lewy body disease (LBD), which includes Parkinson’s disease (PD) dementia and dementia with Lewy bodies (DLB). It affects function and quality of life and is directly associated with reduced survival.^1,2^ OH in the absence of compensatory tachycardia is referred to as *neurogenic* orthostatic hypotension (nOH). It arises from impaired regulation of sympathetic control by central neural pathways or from inadequate activation of vascular adrenoceptors secondary to degenerative postganglionic sympathetic neurons.^3^

CVAD is associated with impaired cognition throughout the disease course in LBD. In prodromal disease, CVAD predicts cognitive impairment.^4^ In PD, autonomic dysfunction caused by dopamine-replacement is associated with worse cognitive fluctuations. This relationship stands even in individuals without cognitive impairment at screening.^5,6^ In advanced DLB, CVAD associates longitudinally with cognitive fluctuations and autonomic burden correlates with neuropsychiatric burden.^7^

The relationship between CVAD and cognition may be primarily explained by impairment in the locus coeruleus (LC), the primary source of noradrenaline. In Alzheimer’s disease, integrity of the LC predicts cognitive function, independent of cortical atrophy.^8^ Imaging the LC and its connections is thought to be a robust biomarker for noradrenergic dysfunction and its effect on neuropsychiatric manifestations in LBD.^9^ Simultaneously, basal forebrain (BF) integrity, through its effect on acetylcholine has been noted to impact cognition in PD, with BF volume mediating the relationship between acetylcholine and delayed memory.^10^ While traditionally considered a feature of Alzheimer’s disease, loss of cholinergic neurons in the BF and cholinergic denervation of the cortex and thalamus has been observed in PD.^11^ Cholinergic lesions were first to be documented in PD^12^, and are thought to be at least as severe as Alzheimer’s disease with a more robust response to acetylcholinesterase inhibitor treatment. Given worse cholinergic markers in those with dementia, the appearance of dementia in PD has been hypothesized to be secondary to cholinergic dysfunction with the BF as a core component.^13,14^

Through this effort, we sought to investigate the LC-BF tract in individuals with LBD and nOH to better understand the underpinnings of the relationship between CVAD and cognitive impairment.

## Materials and Methods

### Study design and participants

Data analyzed was prospectively collected as part of the Cincinnati Cohort Biomarkers Program (CCBP).^15^ The study protocol was approved by the University of Cincinnati Institutional Review Board (IRB# 2018–5699). We performed a cross-sectional, tract-based imaging analysis. The analytic sample included clinical, autonomic, cognitive, and tractography data on 52 participants with a clinical presentation consistent with a diagnosis of PD or DLB. Nine participants had nOH and 43 did not. Cognitive status included 15 participants with normal cognition, 26 with mild cognitive impairment (MCI), and 11 with dementia. (Table 1)

**Table 1.** Baseline clinical and demographic data on study participants.

| Characteristic | No nOH (n=43) | nOH (n=9) | Total (n=52) |
| --- | --- | --- | --- |
| Male sex | 24 (56%) | 8 (89%) | 32 (62%) |
| Race |  |  |  |
| Black or African American | 1 (2%) | 1 (11%) | 2 (4%) |
| White | 42 (98%) | 8 (89%) | 50 (96%) |
| Ethnicity |  |  |  |
| Hispanic or Latino | 1 (2%) | 0 (0%) | 1 (2%) |
| Not Hispanic or Latino | 42 (98%) | 9 (100%) | 51 (98%) |
| Education (years), mean ± SD (minimum, maximum) | 15.9 ± 2.4 (12, 21) | 17.2 ± 3.5 (12, 21) | 16.1 ± 2.6 (12, 21) |
| Age at MRI (years), mean ± SD (minimum, maximum) | 65.9 ± 11.3 (39, 85) | 69.6 ± 5.6 (60, 77) | 66.6 ± 10.6 (39, 85) |
| Age at onset of parkinsonism (years), median (P25, P75) | 61.7 (51.5–68.6) | 60.6 (56.9–63.3) | 61.1 (52.3–66.8) |
| Disease duration (years), median (P25, P75) | 5.3 (2.8–8.8) | 8.2 (7.7–11.7) | 5.9 (3.0–9.8) |
| Cognitive status |  |  |  |
| Unaffected/ normal | 13 (30%) | 2 (22%) | 15 (29%) |
| Mild cognitive impairment | 22 (51%) | 4 (44%) | 26 (50%) |
| Dementia | 8 (19%) | 3 (33%) | 11 (21%) |
| MDS-UPDRS part III score, median (P25, P75) | 23 (16–35) | 29 (18–34) | 23 (16–34) |
| LEDD (mg), median (P25, P75) | 600 (400–980) | 800 (400–1400) | 650 (400–990) |
| Number of anti-hypotensive medications |  |  |  |
| 0 | 43 (100%) | 7 (78%) | 50 (96%) |
| 1 | 0 (0%) | 1 (11%) | 1 (2%) |
| 2 | 0 (0%) | 1 (11%) | 1 (2%) |
| SBP (mm Hg), median (P25, P75) | 130 (118, 146) | 152 (127, 167) | 131 (120, 154) |
| DBP (mm Hg), median (P25, P75) | 82 (75, 90) | 79 (76, 97) | 81 (75, 90) |

Legend: DBP = diastolic blood pressure; LEDD = levodopa equivalent daily dose; MDS-UPDRS = Movement Disorder Society–sponsored revision of the Unified Parkinson's Disease Rating Scale; nOH = neurogenic orthostatic hypotension; P25 = 25th percentile; P75 = 75th percentile; SBP = systolic blood pressure; SD = standard deviation.

Informed consent was obtained from all subjects with conduct consistent with the principles of the Declaration of Helsinki.

### Cognitive status and clinical covariates

Cognitive status was classified as normal cognition, MCI, or dementia. If the individual had a MoCA <21^16^ and cognitive impairment affected ADLs, and/or they were on a medication prescribed with the intent of addressing cognitive impairment such as Donepezil, Rivastigmine or Memantine, they were classified as having dementia. Cognitive impairment or MoCA between 21 and 26^16^, but with no impact on ADLs constituted MCI. For the primary analysis, cognitive status was dichotomized as normal/ unaffected cognition versus cognitive impairment. DLB diagnosed followed the 2017 McKeith criteria.^17^

### Neurogenic orthostatic hypotension (nOH)

nOH was defined by a ratio between the heart rate response to systolic BP reduction of <0.5 beats/min/mmHg with 3-minute of active standing.^18^

### MRI Acquisition

Each participant underwent MRI on a GE 3T Signa Architect scanner consisting of T1-weighted Brain Volume imaging (T1 BRAVO), T2-weighted Cube imaging (T2), T2 Fluid Attenuated Inversion Recovery Cube imaging (T2 FLAIR), Susceptibility Weighted Imaging (SWAN), high-directional multi-shell diffusion MRI (dMRI), and resting state functional MRI (rs-fMRI). Only the T1 BRAVO (TR/TE=7.69/3.12ms, TI=450ms, Flip Angle = 12, slice thickness = 1mm, FOV = 25.6cm, Matrix = 256×256, resolution = 1×1×1mm3), T2 (TR/TE=3000/100ms, Flip Angle = 90, slice thickness = 1mm, FOV = 25.6cm, Matrix = 256×256, resolution = 1×1×1mm3), and dMRI (b=1000/2000/3000, directions=7/29/64, 3 b0, TR/TE=7134/97.2ms, FOV=24.0, Matrix=96×96, slice thickness = 2.5mm, resolution = 2.5×2.5×2.5mm3, hyperband = 3) were used for this analysis.

### MRI Preprocessing

T1 and T2 preprocessing included bias field correction and brain extraction using HD-BET. dMRI data were preprocessed using TORTOISE, including eddy-current distortion correction, T2-based geometric distortion correction, Gibbs ringing correction, denoising, and motion correction, with rotation of gradient directions where appropriate.

### dMRI Tract-Specific Analysis

After preprocessing, dMRI data were reconstructed using generalized q-sampling imaging (GQI). The GQI data for each participant were used to generate participant-specific tractography to investigate differences in the ventral ascending reticular activating system (vARAS) traversing the LC and BF (Figure 1). LC and BF regions of interest (ROIs) in MNI-152 template space were mapped to participant diffusion space. Briefly, a brain-extracted T1 MNI-space template was nonlinearly warped to each participant’s T1 BRAVO using ANTs; the T1 BRAVO was then aligned through the T2 image to the preprocessed dMRI data, and the resulting transforms were applied to the ROIs. Deterministic tractography based on quantitative anisotropy (QA) was performed between aligned ROIs to reconstruct the LC-BF tract. Tracking parameters were: normalized QA threshold = 0.2, turning angle = 60, step size = 1.0, minimum length = 20mm, maximum length = 100mm, and number of tracks = 10,000. A region of avoidance was placed in the cerebellum to minimize false-positive fibers. Each resulting tract was reviewed by three trained experts (BW, KD and DR) and manually cleaned to remove false-positive fibers when needed. Raters were blinded to nOH status and all other clinical information.

**Figure 1.**
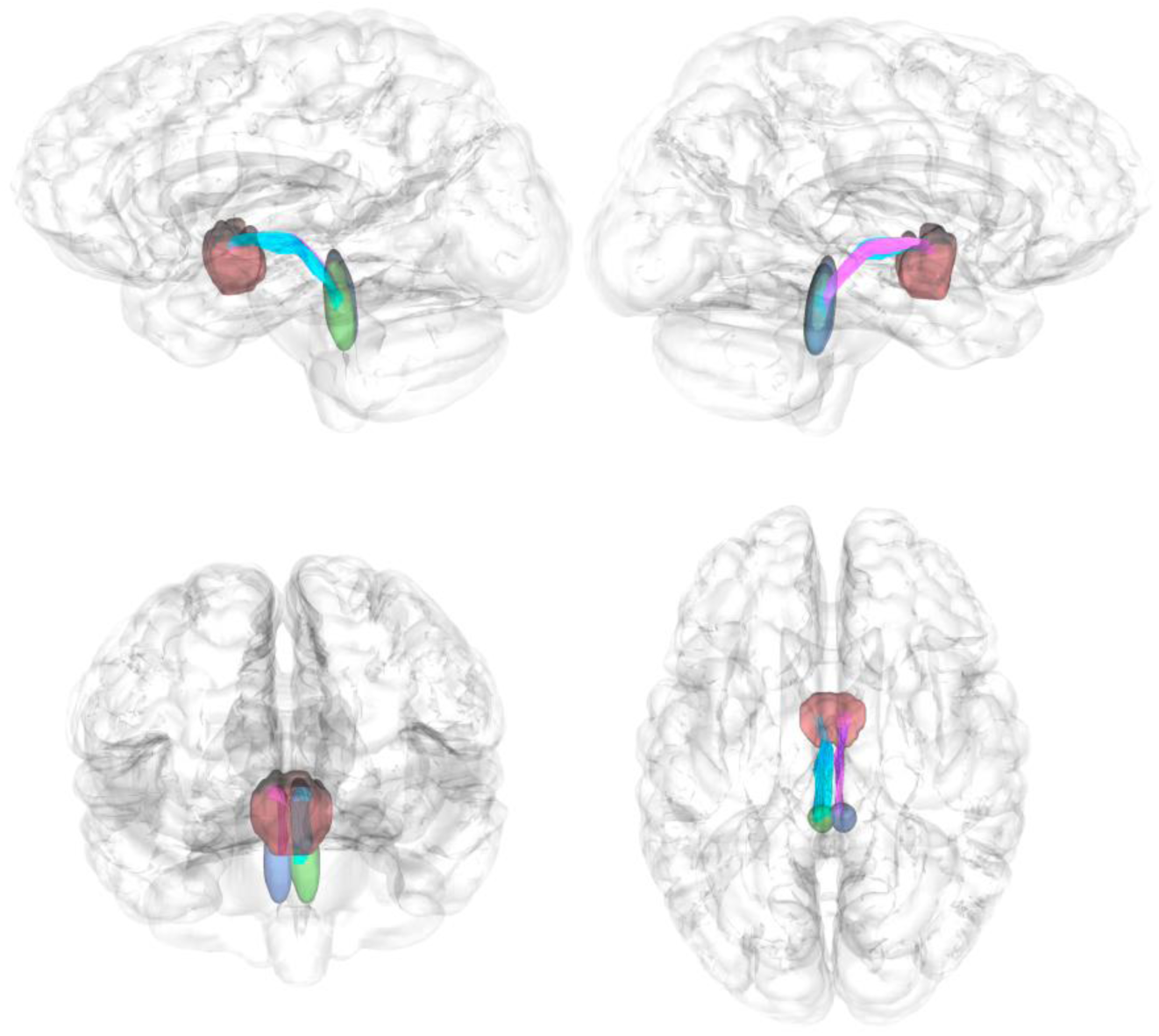
LC-BF tract reconstruction and regional macrostructural quantification. Representative bilateral locus coeruleus-basal forebrain (LC-BF) tract reconstructions are shown with regions-of-interest (ROIs), which included left LC (green), right LC (blue), and basal forebrain (red). Macrostructural endpoints included total tract volume, total surface area, and segmental tract measures, including terminal fourth-quarter tract volume. LC-BF = locus coeruleus-basal forebrain.

### Tract characteristics

Primary macrostructural tract characteristics included total volume, first-quarter volume, fourth-quarter volume, end-branch volumes, and total surface area. Microstructural measures included quantitative anisotropy (QA), isotropic diffusion (ISO), radial diffusivity (RD), and QA-to-isotropic diffusion ratio (QIR). Left and right LC-BF tract metrics were averaged to create bilateral tract metrics.

### Primary statistical analysis

The primary analysis used Bayesian semiparametric proportional-odds models implemented with the blrm function from the rmsb package in R. Continuous tract metrics were analyzed as ordered responses under the proportional-odds framework; therefore, odds ratios below 1 indicate a lower adjusted tendency toward higher tract-metric values among participants with nOH compared with participants without nOH. For each tract metric, the primary model evaluated the association between nOH and LC-BF tract characteristics with adjustment for age at MRI, sex, and education. Age at MRI was modeled using restricted cubic splines with three knots to allow for nonlinear age associations. Default priors were used with four Markov chains with 4,000 iterations per chain, including 1,000 warmup iterations.

### Sensitivity and secondary analyses

Sensitivity analyses evaluated the consistency of primary nOH associations after additional adjustment for ordinal cognitive severity, high blood pressure, disease duration to MRI, and combined high blood pressure plus disease duration. Secondary cognition-focused analyses tested associations between cognitive status and prespecified LC-BF tract endpoints using a binary normal cognition versus MCI/dementia contrast adjusted for age at MRI, sex, education, and nOH status; ordinal cognitive severity was evaluated as a sensitivity analysis. Volume-adjusted models were used for microstructural endpoints where appropriate. Because multiple correlated tract endpoints were evaluated, inference emphasized posterior effect estimates, 95% credible intervals, posterior directional probabilities, anatomical coherence across related endpoints, and consistency across prespecified models rather than p-values or formal multiplicity correction.

### Model diagnostics

Markov Chain Monte Carlo (MCMC) diagnostics were reviewed for the primary and sensitivity Bayesian models, including divergent transitions, treedepth, effective sample size, split R-hat, and the energy Bayesian fraction of missing information. Results were considered interpretable when models completed successfully, contrast estimates were extractable, and diagnostics did not suggest major sampling problems.

## Results

### Participant characteristics

The analytic cohort included 52 participants, of whom 9 had nOH and 43 did not. The nOH distribution within cognitive strata was 2 of 15 participants with normal cognition, 4 of 26 with MCI, and 3 of 11 with dementia. A full description of the cohort is shown in Table 1.

### Model diagnostics

Across all models (primary and sensitivity), MCMC diagnostics did not identify major convergence or sampling concerns. There were no divergent transitions, maximum treedepth was 5, maximum split R-hat was 1.003, minimum bulk effective sample size was 13,958, minimum tail effective sample size was 6,600, and minimum energy Bayesian fraction of missing information was 0.803.

### Macrostructural Tract Metrics

In the demographic-adjusted model, nOH was associated with lower LC-BF bilateral fourth-quarter volume (OR = 0.19, 95% CrI 0.04–0.81; Pr[OR < 1] = 0.987), left fourth-quarter volume (OR = 0.13, 95% CrI 0.03–0.58; Pr[OR < 1] = 0.997), left total surface area (OR = 0.19, 95% CrI 0.04–0.77; Pr[OR < 1] = 0.990), and left total volume (OR = 0.22, 95% CrI 0.05–0.91; Pr[OR < 1] = 0.982). These associations were directionally consistent and of similar magnitude after additional adjustment for cognitive severity, high blood pressure, disease duration, and combined high blood pressure plus disease duration (Figure 2). A representative example demonstrating these macrostructural differences is shown in Figure 3.

**Figure 2.**
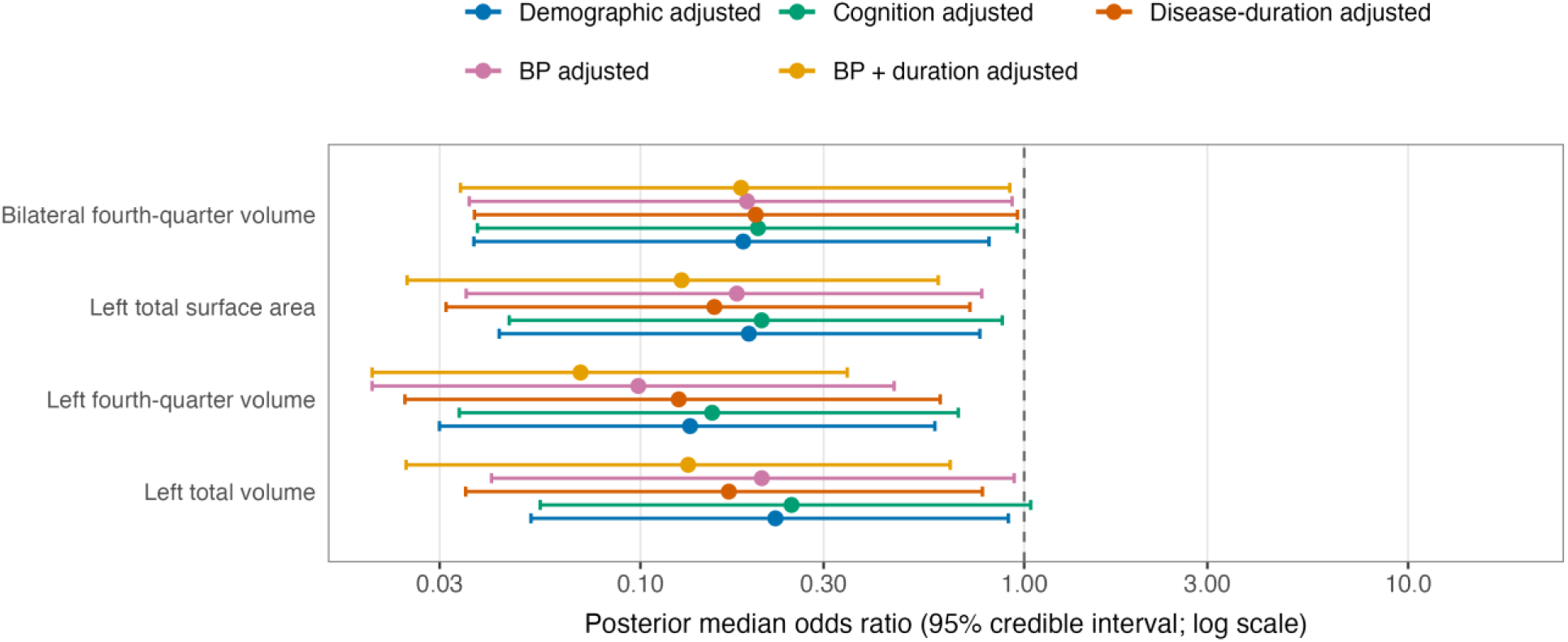
Neurogenic orthostatic hypotension shows consistent associations with lower LC-BF macrostructural tract measures across covariate-adjusted models. Forest plot showing posterior median odds ratios and 95% credible intervals for associations between nOH and selected LC-BF macrostructural endpoints in the disease-only neurodegenerative cohort. Models included demographic adjustment, additional adjustment for ordinal cognitive severity, additional adjustment for disease duration, high blood pressure adjustment, and combined high blood pressure plus disease-duration adjustment. Odds ratios less than 1 indicate a lower tendency toward higher tract-metric values among participants with nOH. Across models, nOH showed consistent associations with lower left-sided and terminal LC-BF macrostructural measures, particularly left fourth-quarter volume, bilateral fourth-quarter volume, left total surface area, and left total volume.

**Figure 3.**
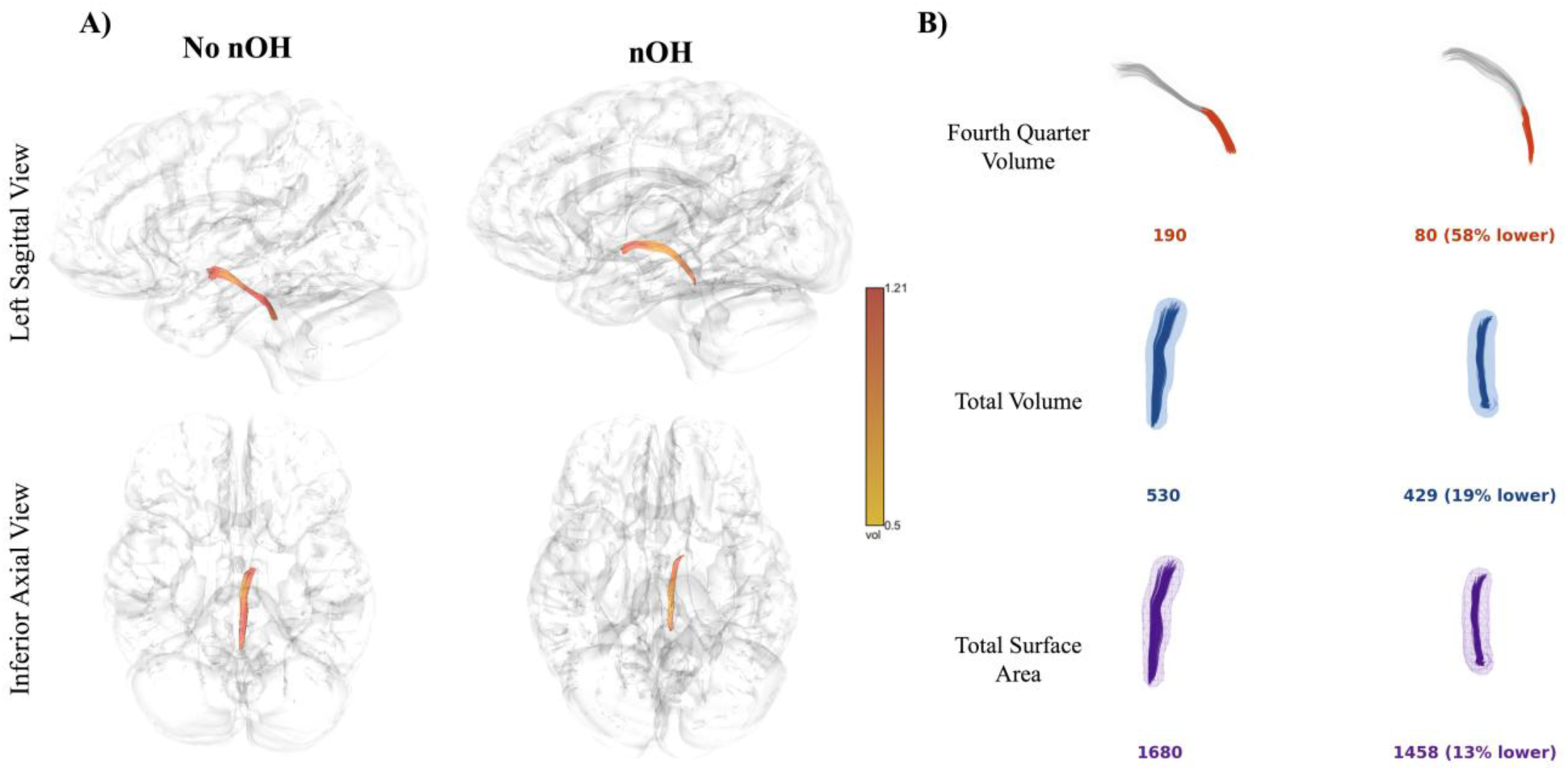
Representative LC-BF tractography visualization of nOH-associated macrostructural differences. **A)** Representative left LC–BF tract reconstructions are shown for participants without nOH and with nOH in sagittal and inferior axial views, providing anatomical context for the tractography-based analyses. Tracts are overlaid on semi-transparent brain renderings and color-coded by tract volume density. **B)** Side-by-side visualizations of tract-derived macrostructural endpoints are shown for the same representative participants, including fourth-quarter volume, total tract volume, and total surface area. The nOH patient demonstrates lower fourth-quarter volume, total volume, and surface area relative to the no nOH example, consistent with the macrostructural endpoints highlighted in the primary models. Values below each rendering indicate the tract-derived metric for that participant, with percent difference shown for the nOH example. These renderings are illustrative examples intended to aid interpretation of the model-based findings and are not independent statistical maps.

### Microstructural Tract Metrics

nOH-related microstructural associations were weaker and less consistent than macrostructural associations. Directional patterns were observed, including lower left QA (OR = 0.48, 95% CrI 0.11–2.06; Pr[OR < 1] = 0.834) and higher left RD (OR = 1.58, 95% CrI 0.38–6.68; Pr[OR > 1] = 0.730), but posterior directional probabilities were weaker than those observed for macrostructural endpoints and did not show comparable consistency across sensitivity models. Volume-related sensitivity analyses did not suggest that the weaker microstructural patterns were attributable to tract volume (left QA: OR = 0.46, 95% CrI 0.09–2.31; Pr[OR < 1] = 0.829; left RD: OR = 1.58, 95% CrI 0.33–7.93; Pr[OR > 1] = 0.722).

### Cognitive status and LC-BF tract characteristics

After adjustment for age, sex, education, and nOH status, participants with impaired cognition showed lower bilateral fourth-quarter volume (OR = 0.33, 95% CrI 0.09–1.09; Pr[OR < 1] = 0.964), higher bilateral QA (OR = 3.13, 95% CrI 0.93–10.68; Pr[OR > 1] = 0.967), and higher bilateral ISO (OR = 2.80, 95% CrI 0.84–9.47; Pr[OR > 1] = 0.953) (Figure 4). These findings suggest that cognitive impairment was associated with lower terminal LC-BF tract volume and higher microstructural metrics related to both axonal density (QA) and edema/gliosis (ISO).

**Figure 4.**
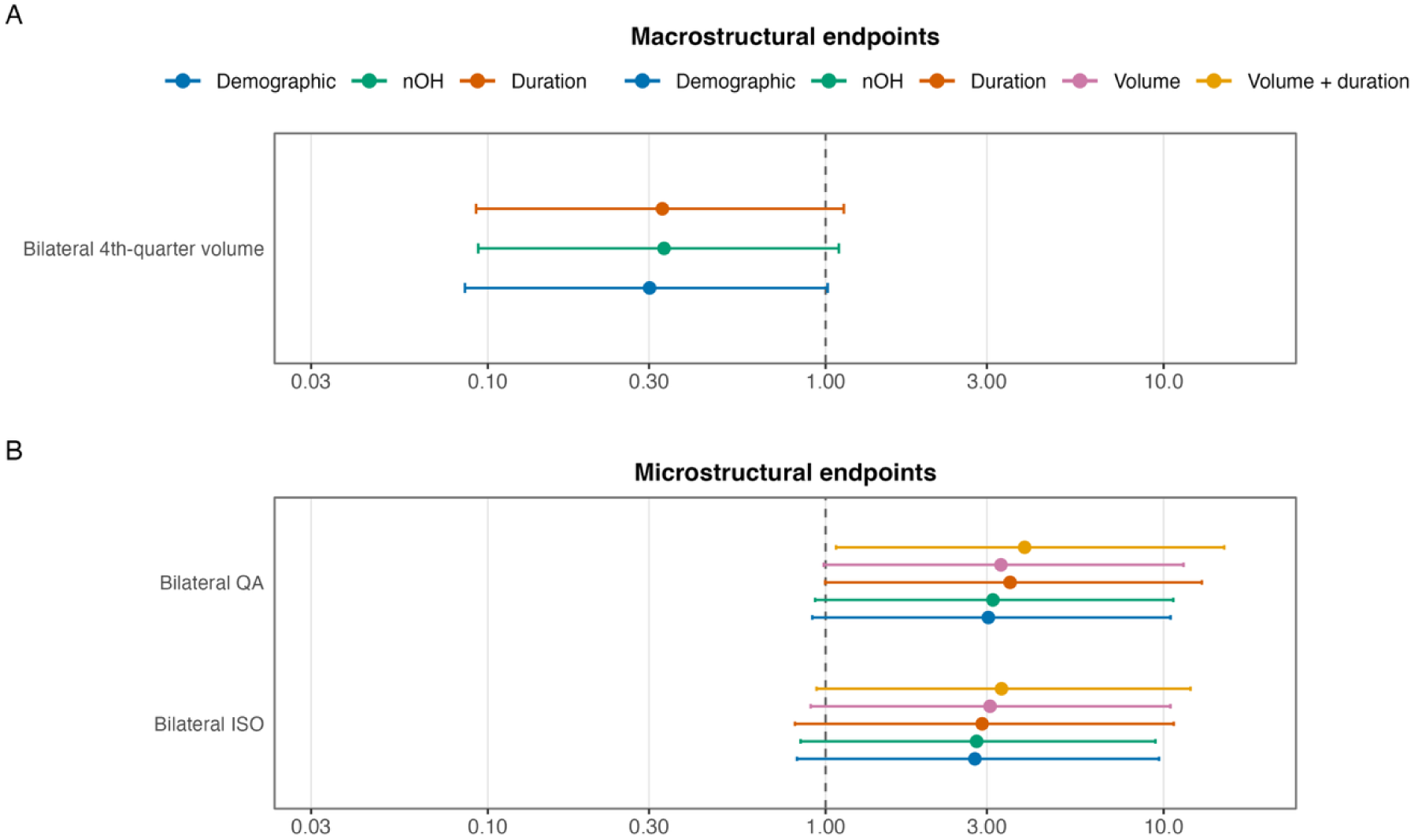
Binary cognitive-status associations with selected LC-BF tract endpoints. Forest plots show posterior median odds ratios and 95% credible intervals for LC-BF tract endpoints associated with binary cognitive status, comparing participants with MCI/dementia to cognitively normal participants. Displayed endpoints met the prespecified posterior directional-probability threshold of 0.95 in the nOH-adjusted model. **A)** Macrostructural endpoints show lower bilateral fourth-quarter volume in participants with MCI/dementia. **B)** Microstructural endpoints showing higher bilateral QA and bilateral ISO in participants with MCI/dementia. Models included demographic adjustment, nOH adjustment, disease-duration adjustment, and, for microstructural endpoints, volume-adjusted and volume-plus- duration-adjusted sensitivity models. Odds ratios less than 1 indicate lower tendency toward higher tract-metric values in MCI/dementia, whereas odds ratios greater than 1 indicate higher tendency toward higher tract-metric values in MCI/dementia. LC-BF = locus coeruleus–basal forebrain; MCI = mild cognitive impairment; nOH = neurogenic orthostatic hypotension; QA = quantitative anisotropy; ISO = isotropic diffusion.

In the sensitivity models investigating the relationship between ordinal cognitive severity and LC-BF tract characteristics, Left ISO was higher with increasing cognitive impairment severity (OR = 2.35, 95% CrI 0.94–5.79; Pr[OR > 1] = 0.967). Other prespecified macrostructural and microstructural endpoints, including quarter-volume measures, total volume, total surface area, QA, RD, and QIR, did not meet the 0.95 directional probability threshold.

## Discussion

Neurogenic orthostatic hypotension consistently associates with lower LC-BF volume, and this volume inversely correlates with the severity of cognitive impairment. Noradrenergic projections from the LC to the forebrain act through alpha and beta adrenoreceptors. These projections may affect cognition and are impacted by neurodegeneration.^19^ LC, through release of norepinephrine, is central to modulating attention, emotion, and regulation of the stress response. LC norepinephrine levels are reduced in PD patients with dementia compared with those without.^19^ Though without strict boundaries, BF is used to describe the region encompassing substantia innominate, portions of the hypothalamus and the ventral striatum with nucleus accumbens.^20^ It includes the basal nucleus of Meynert, with neurons rich in the neurotransmitter, acetylcholine. In individuals with PD dementia, the basal nucleus of Meynert suffers a 60-65% reduction of choline acetyltransferase positive neurons coupled with extensive cholinergic fiber degeneration.^21^ BF volume reduction in those with PD-MCI predicts progression to dementia.^22^ LC-BF tract volume robustly associates with cognition in LBD, highlighting the mechanistic underpinnings behind the relationship seen between CVAD and cognition.

Macrostructural measures reflect fiber volume and can therefore be used to detect fiber loss or tract atrophy, while microstructural metrics represent integrity of the non-atrophied fibers. In LBD, it is likely that degeneration of the LC leads to fewer noradrenergic projects, leading to low tract volume. Our results revealed no differences in microstructural metrics between those with and without nOH. Volume-adjusted sensitivity analyses conducted to ensure that microstructure findings were not simply collinear or explained by volume did not change these results. In addition, correction of the microstructure by number of streamlines did not explain the differences in macrostructural and microstructural measures. Such differences may be hypothetically explained by relative preservation of diffusion properties by the remaining axons, gliosis or predominant gray matter neuron loss^23^, leading to no noted difference in the measured tract integrity with nOH. Based on these results, LC-BF tract morphology is a more sensitive biomarker of nOH in LBD than diffusion metrics.

Better detection and understanding of the pathophysiology behind nOH-associated cognitive impairment has implications beyond clinical presentation. A study of neuropsychological testing in the upright and supine positions in individuals with PD revealed that results may vary depending on the presence of nOH.^24^ Benefit seen with Rivastigmine, the only FDA-approved medication for treatment of PD dementia, is greater in those with OH and may partly be secondary to an anti-hypotensive effect.^25^

This study has several limitations. nOH was defined from the ratio of heart-rate response to blood pressure reduction during 3-minute active standing. This provides a clinically relevant marker of neurogenic orthostatic physiology but does not capture the cumulative burden of blood pressure instability over time, including recurrent hypotensive episodes, supine hypertension, post-prandial hypotension, or medication-related blood pressure fluctuations. Cognitive status was analyzed using clinical categories of normal cognition, MCI, and dementia. These categories are clinically meaningful but do not capture domain-specific cognitive performance, cognitive fluctuations, attentional variability, or longitudinal cognitive decline. This is relevant because LC-BF pathways may be particularly related to arousal, attention, and fluctuation-related symptoms rather than global cognitive status alone. The cohort reflects the clinical population in which nOH and cognitive impairment commonly overlap. While these observations are likely seen across clinical diagnoses, the distribution of autonomic burden, LC-BF involvement, disease duration, and medication exposure may vary. Studies with a larger sample size could help determine whether the observed LC-BF associations differ across PD, PD dementia, DLB, and related parkinsonian syndromes. It may additionally reduce the 95% confidence interval, lending even greater certainty to the results. Finally, tractography-derived measures remain indirect imaging markers. Tract volume, surface area, and fourth-quarter volume reflect the extent of the reconstructed LC-BF pathway rather than direct measures of neuronal or axonal loss. Similarly, QA, ISO, RD, and QIR reflect diffusion properties of the reconstructed pathway and surrounding tissue environment and should not be interpreted as simple one-to-one measures of tract integrity.

### Future directions

Such imaging of brain structural alterations, including those associated with neurodegeneration, provides a framework for predicting cognitive losses in LBD. These results must be tested in a larger cohort to determine if they are generalizable. Future efforts should investigate the utility of these results to help identify high-risk individuals for more precise prognostication and earlier intervention, including enrollment in clinical trials. While commonly prescribed for a number of symptoms, the therapeutic potential of noradrenergic therapies has not been yet realized.^26,27^ Appropriate investigation of such a biomarker may characterize the patient population likely to benefit from novel therapeutic avenues for individuals with LBD.

## Data Availability

Data analyzed as part of this effort will be available on reasonable request.

## Acknowledgements

We want to thank all subjects who participated in the CCBP study.

